# Comparative experimental evidence on compliance with social distancing during the COVID-19 pandemic

**DOI:** 10.1101/2020.07.29.20164806

**Authors:** Michael Becher, Daniel Stegmueller, Sylvain Brouard, Eric Kerrouche

**Affiliations:** Institute for Advanced Study in Toulouse and IE School of Global and Public Affairs; Duke University; CEVIPOF, Science Po, Paris

## Abstract

Social distancing is a central public health measure in the fight against the COVID-19 pandemic, but individuals’ compliance cannot be taken for granted. We use a survey experiment to examine the prevalence of non-compliance with social distancing in nine countries and test pre-registered hypotheses about individual-level characteristics associated with less social distancing. Leveraging a list experiment to control for social desirability bias, we find large cross-national variation in adherence to social distancing guidelines. Compliance varies systematically with COVID-19 fatalities and the strictness of lockdown measures. We also find substantial heterogeneity in the role of individual-level predictors. While there is an ideological gap in social distancing in the US and New Zealand, this is not the case in European countries. Taken together, our results suggest caution when trying to model pandemic health policies on other countries’ experiences. Behavioral interventions targeted towards specific demographics that work in one context might fail in another.

## Introduction

In the fight against epidemics, including the novel coronavirus disease (COVID-19) caused by the Sars-Cov2 virus, large-scale behavioral change is essential to limit the loss of human lives and to allow societies to resume economic and social activities. After the outbreak of COVID-19 in 2019 and its global spread as a pandemic in the first half of 2020, the absence of vaccination and medical treatment meant that non-pharmaceutical interventions—such as social distancing and hand washing—were crucial to mitigate and contain the spread of the virus. Most governments have adopted clear recommendations and rules to limit physical and social contact, and social scientists have immediately started to study people’s compliance with the new behavioral rules. Surveys on COVID-19 in different countries have consistently shown very high rates of self-reported compliance with recommended health norms in the population (Barari et al. 2020; Brouard et al. 2020; Perrotta et al. 2020; Utych and Fowler 2020).

However, several scholars have cautioned that direct survey questions are likely to suffer from measurement error due to social desirability bias (Barari et al. 2020; Daoust et al. 2020). For example, survey research on self-reported behavior and attitudes has shown that survey self-reports of voter turnout or racial animus are affected by the pressure to provide what is perceived as the socially desirably rather than the factually correct answer (e.g., Belli et al. 2001; Bernstein et al. 2001; Kuklinski et al. 1997). While some scholars have reported that online-mode surveys reduce the impact of desirability bias (Holbrook and Krosnick 2010), the public salience of these health measures may still induce overreporting of compliant behavior (Barari et al. 2020: 4; Munzert and Selb 2020). Thus, social desirability is likely to be a factor when respondents are directly asked to report whether they complied with highly-publicized behavioral rules during a pandemic, when non-compliance is depicted as irresponsibly putting the lives of others at risk. Misreporting of behavior makes it more difficult to identify the groups that are least likely to comply and could be targeted for further interventions (Bavel et al. 2020; West et al. 2020).

To mitigate this measurement problem and to provide more robust insights on the determinants of non-compliance with social distancing during the COVID-19 pandemic, we use a list experiment (Miller 1984; Raghavarao and Federer 1979) as a measurement device designed to reduce social desirability bias. We embedded it into a comparative internet survey covering eight countries in lockdown in mid-April 2020 (Australia, Austria, France, Germany, Italy, New Zealand, United States, United Kingdom) as well as Sweden. The latter took a less stringent policy response, but still recommended social distancing. The list experiment (also called unmatched or item count technique) allows respondents to truthfully report their behavior with respect to social distancing without revealing it to the researcher.

Faced with a list of items, respondents are asked how many of these things they have done last week, but not which specific ones. Respondents are randomly assigned to treatment and control groups. The treated group received an additional item capturing the violation of the social distancing norm. The key assumption is that the treatment group would have responded like the control group absent the treatment. Importantly, the list experimental design can be leveraged to study individual-level correlates of not adhering to the norm (Imai 2011; Blair and Imai 2012). When using these results to guide public policy, this provides a clear advantage over (anonymized) data from smartphones or credit card transaction, which require geo-spatially aggregated variables of interest (Allcott et al. 2020; Painter and Qiu 2020). Using aggregates to infer the behavior of individuals poses the risk of ecological fallacies (Robinson 1950; Greenland and Robins 1994).

In this paper we provide comparative experimental estimates of non-compliance with social distancing by meeting friends or relatives. Our sample of nine advanced industrialized democracies covers approximately 65% of total confirmed COVID-19 related deaths at the time of the survey (Dong et al. 2020). It covers large variation in mortality (from less than five deaths per million inhabitants in Australia and New Zealand to more than 350 in Italy) and governmental responses (from very strong restrictions in Italy, New Zealand and France to comparatively few restrictions in Sweden).^1^

Our analysis yields two main sets of results. First, it reveals a substantial degree of non-adherence to social distancing guidelines. In most countries under study, experimental estimates of non-compliance are much higher than estimates based on direct questions from other surveys fielded in the same countries at the same time, which offer a more optimistic picture (Perrotta et al. 2020). Moreover, we find that cross-national variation in rates of non-compliance is negatively correlated with the severity of the crisis, proxied using confirmed COVID-19 related deaths, as well as the stringency of lockdown-style policies that limit people’s movement and social activities.

Second, our analysis provides new insight into individual-level characteristics associated with non-compliance in each country. Achieving compliance with collective decisions is a general problem that states tackle with mix of monitoring, sanctions and voluntary cooperation. A large and cross-disciplinary literature on compliance suggests that for a given level of external enforcement, individuals may vary in their willingness to adapt their behavior to COVID-19 guidelines (Levi and Stoker 2000; Luttmer and Singhal 2014). Beyond socio-demographic variables (age, gender, and education), which have received most attention in studies of compliance with social distancing in particular and non-pharmaceutical interventions in general, we test pre-registered hypotheses concerning the role of political ideology and trust. Our results uncover substantial heterogeneity among countries. In the US and in New Zealand, we find that there is an ideological gap in social distancing. People that place themselves on the extreme right of the political spectrum are less likely to practice social distancing than those with centrist views, whereas people with extreme left beliefs are more likely to comply with social distancing. These results highlight important political constraints in the fight against the pandemic. They are also consistent with recent evidence from the US on the partisan gap in compliance between Democrats and Republicans (Allcott et al. 2020; Green et al. 2020; Painter and Qiu 2020). By contrast, in most European countries under study political ideology is not linked to compliance with social distancing. The role of trust also varies by country. Being more trusting of others is associated with more social distancing in some countries (Germany), consistent with theories of social dilemmas and public good provision, less social distancing in others (US), and not associated in others.

These results have two broader implications for policy choices during the pandemic. First, the heterogeneity of individual-level results across countries suggests that it may be difficult to learn from other countries’ experiences. Identifying the characteristics of non-compliers, who could be targeted or nudged into more compliance, might be a task that depends on country-specific idiosyncrasies unlikely to be guided well by using results from other countries. Second, social distancing appears to be more difficult to maintain when the most severe restrictions and external sanctions have been lifted and mortality rates are declining.

## Empirical Results

In this section we present our empirical findings. Section ‘Materials and Methods’ provides more details about the survey and its context, the statistical methods used to analyze the experiment and validity checks. As noted in the introduction, the list experiment asks respondents how many things—not which ones—from a list of items they have done last week. The control group received a list of four behaviors that are generally permissible under existing health recommendations (such as ordering food using online delivery services or seeking medical care; see Online Appendix (OA) A.2 for the full list). In addition to these four items, the treatment group received a more sensitive item that indicates a lack of social distancing: “I met with two or more friends or relatives who do not live with me.” Given this design, the difference-in-means between between the item count in the treatment group and the control group, weighted by the sampling probabilities, estimates the prevalence of not following social distancing in the population.

### Country-level non-compliance with social distancing

Table I shows the estimated fraction of people in each country who met two or more friends or relatives not living in their household during the previous week. The estimates reveal a substantial degree of non-adherence to social distancing guidelines during the pandemic. In six out of eight countries a large and statistically significant fraction of the population did not follow social distancing guidelines. In some countries under lockdown (Austria and Germany), a (near-)majority of the population met friends or relatives despite the explicit health recommendations against it. In the US and Australia, a large minority (of at least 20% or more) of the population did not follow the norm. Experimental estimates of non-compliance are lower (but still statistically significantly different from zero) for France (13%) and New Zealand (12%). The fraction of non-compliers is not statistically distinguishable from zero in Italy and the UK. Finally, in Sweden, which did not enact a lockdown and where social distancing recommendations were less strict, around half of the population (48%) met friends or relatives.

**Table I.**
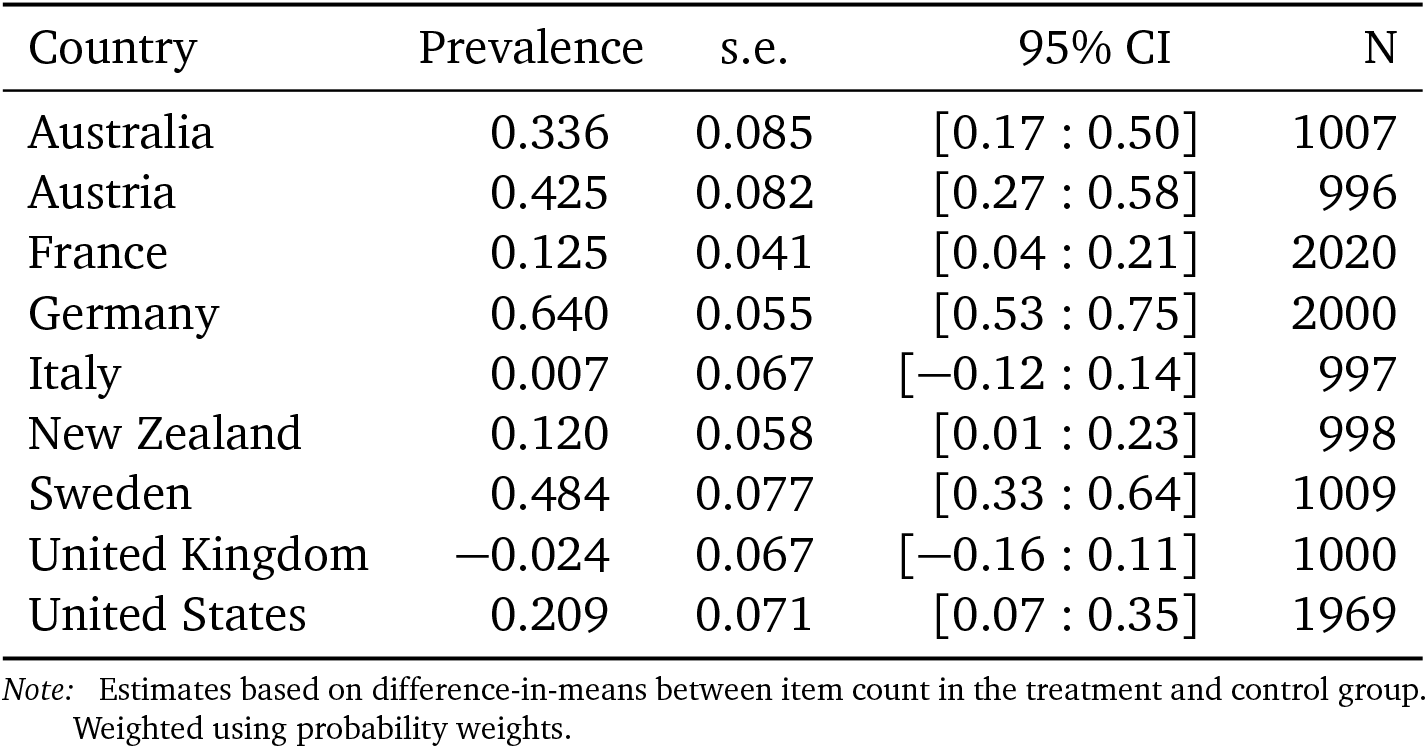
**Experimental estimates of prevalence of individuals not following social distancing guidelines during the COVID-19 pandemic in 9 countries**.

| Country | Prevalence | s.e. | 95% CI | N |
| --- | --- | --- | --- | --- |
| Australia | 0.336 | 0.085 | [0.17 : 0.50] | 1007 |
| Austria | 0.425 | 0.082 | [0.27 : 0.58] | 996 |
| France | 0.125 | 0.041 | [0.04 : 0.21] | 2020 |
| Germany | 0.640 | 0.055 | [0.53 : 0.75] | 2000 |
| Italy | 0.007 | 0.067 | [−0.12 : 0.14] | 997 |
| New Zealand | 0.120 | 0.058 | [0.01 : 0.23] | 998 |
| Sweden | 0.484 | 0.077 | [0.33 : 0.64] | 1009 |
| United Kingdom | −0.024 | 0.067 | [−0.16 : 0.11] | 1000 |
| United States | 0.209 | 0.071 | [0.07 : 0.35] | 1969 |

*Note:* Estimates based on difference-in-means between item count in the treatment and control group. Weighted using probability weights.

### Relationship between non-compliance and COVID-19 severity

Table I revealed substantial variation in adherence to social distancing across countries. While not part of our initial pre-analysis plan, we examine if this variation is systematically related to observable characteristics of the pandemic. The exploratory analyses summarized in Figure I provide two empirical insights. As shown in panel A of Figure I, we find that the estimated share of individuals meeting family and friends is negatively correlated with the total COVID-19 related deaths in the week prior to the survey (per million inhabitants). The data we use are official government-reported counts compiled by researchers at Johns Hopkins University (Dong et al. 2020).^2^ The estimated slope of a robust regression line (section Materials and Methods provides more details on the statistical model) is − 0.011 (with a *p*-value *<* 0.001) and suggests that, on average, countries with lower reported deaths, like Austria or the US, exhibit significantly higher levels of non-compliance than countries with higher reported deaths, such as France and Italy. The bivariate regression describes cases like New Zealand (low non-compliance despite few deaths) or Germany (highest rate of non-compliance) less well.

**Figure I.**
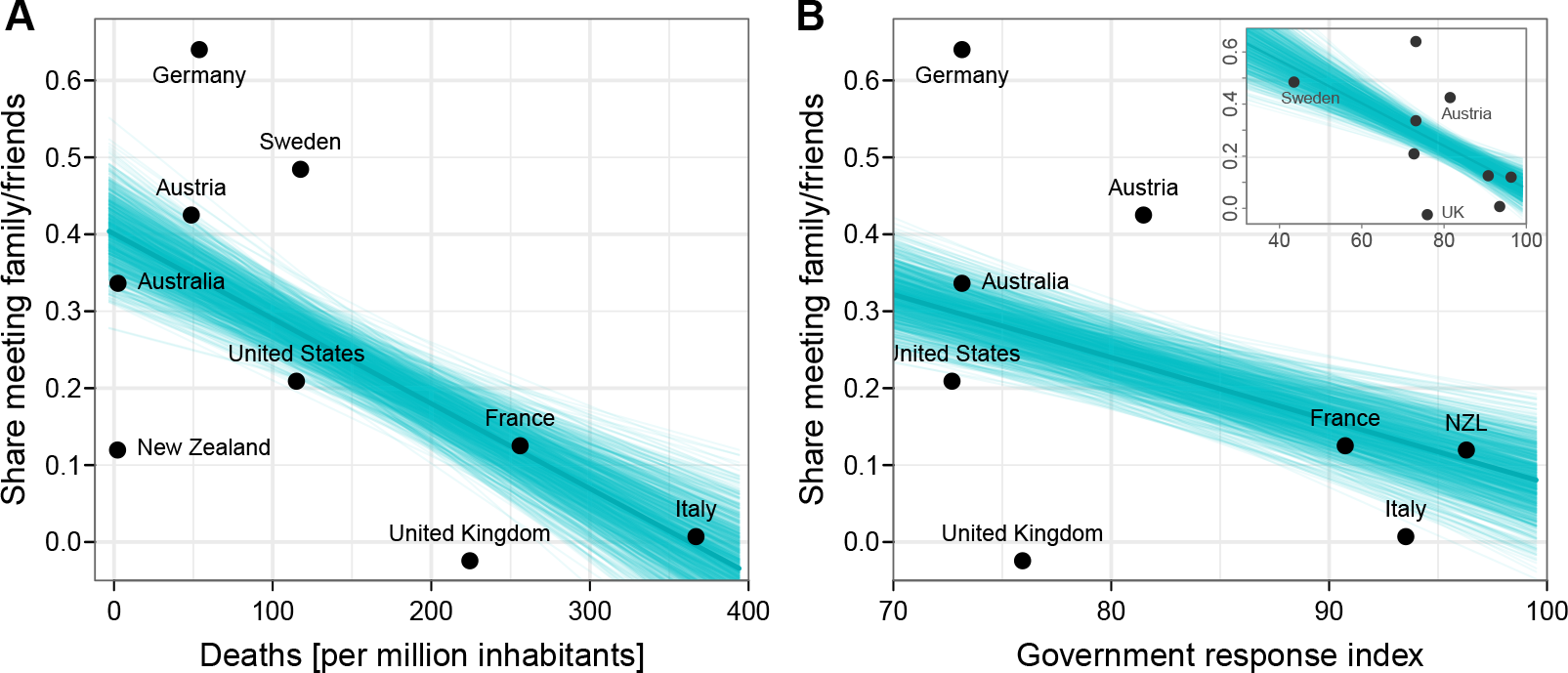
Relationship between population violating social distancing guidelines and COVID-19 related deaths and strictness of lockdown measures. This figure plots the fraction of individuals meeting family and friends estimated from the list experiment (y-axis) against total COVID-19 related deaths in the week before the survey (**A**) and the strictness of lockdown-style measures (**B**). Each plot includes the linear fit from a robust regression (using an M-estimator with Huber objective function) estimated on 1,000 replicate data sets capturing variation in experimental estimates. The inset in panel B shows the plot with an enlarged x-axis including Sweden, which enacted the least stringent measures.

Panel B of Figure I shows a similar negative association between meeting friends and family and the stringency of the government response to the pandemic (the slope of the bivariate regression is − 0.082; *p <* 0.001). This finding is consistent with country-level results from the US (Painter and Qiu 2020). The stringency index is taken from the Oxford COVID-19 Government Response Tracker (Hale et al. 2020). It measures (on a scale from 0 to 100) the strictness of lockdown-style policies, such as restrictions of movement and school closures, that primarily restrict people’s behavior.^3^ In this analysis, New Zealand, where overall restrictions were very high, is no longer an outlier.

The associations displayed in Figure I are meant to illustrate that aggregated individual behavior corresponds to central characteristics of the pandemic and the responses of governments dealing with it. They are not causal statements on macro-micro effects (and the limited number of countries limits the ability to control for possible confounders). Nonetheless, they are consistent with the fundamental idea in the compliance literature that the extent of external monitoring and sanctions are relevant factors (Luttmer and Singhal 2014). It also appears that the severity of the public health crisis likely shapes the salience of the issue and it may also enhance self-interested motivations to follow social distancing. The country patterns of physically meeting friends and relatives during the pandemic are also not simply a product of the intensity of existing social ties in a country. OA Figure A.1 shows that our list experimental estimates have no substantive or statistically significant relationship with the average amount of time spent socializing with friends and family before the pandemic.

### Individual-level predictors of non-compliance

Our second set of empirical estimates examines individual-level predictors of non-compliance with social distancing in each country.^4^ To better isolate the contribution of individual-level factors from macro-level characteristics, such as variation in policies, institutions or the severity of the pandemic, we conduct analyses separately for each country. Figure II displays the estimated relationship between selected demographic and personality characteristics and the probability of meeting friends and relatives despite social distancing guidelines for each country. It displays maximum likelihood estimates (Imai 2011; Blair and Imai 2012) of the change in predicted probability of non-compliance (with 90% confidence intervals) arising from a change in an individual characteristic. We provide two specifications: one studying the bivariate or unadjusted relationship between individual characteristics and the probability of non-compliance, and an adjusted specification that includes a set of individual-level controls capturing some possible mechanisms or alternative explanations (see Materials and Methods for model details). Our estimates reveal considerable cross-national variation in the direction of individual characteristics for social distancing.

The literature on compliance with government decisions shows that political beliefs may be relevant (Levi and Stoker 2000). If this is true in the case of COVID-19 as well, it indicates a considerable challenge for democratic governments trying to encourage compliance with non-pharmaceutical public health measures. Political beliefs are not easily changed and targeting interventions (e.g., messaging or surveillance) to different political groups raises important normative questions. In the US, we find that people who self-identify (on an 11-point ideological scale) as being further on the right are more likely to meet friends or relatives during the pandemic compared to people with more centrist views. Substantively, experimental estimates suggests that individuals one standard deviation to the right of the national ideological mean are approximately 12% more likely to skirt social distancing. In contrast, individuals one standard deviation to the left are approximately 10% more likely to follow social distancing guidelines.

**Figure II.**
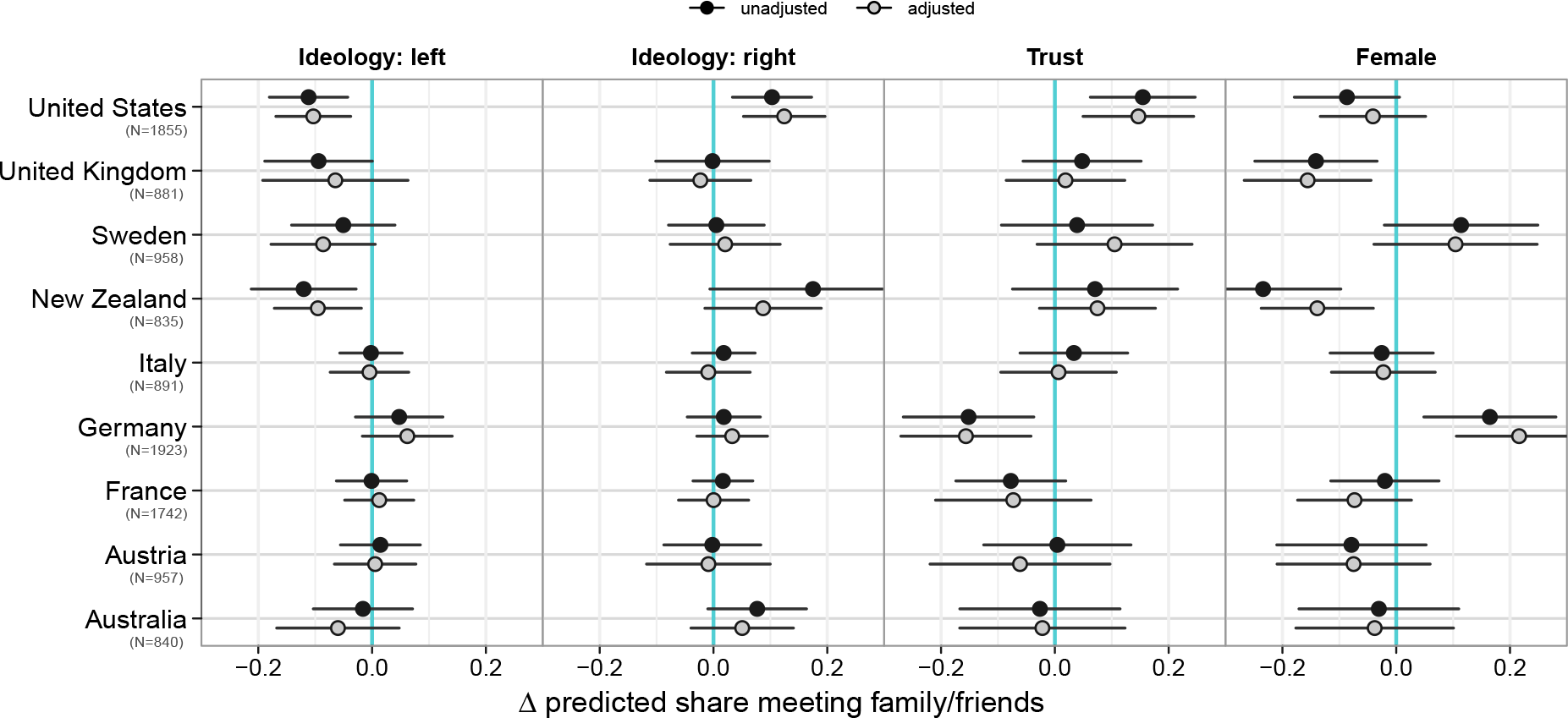
Relationship between individual characteristics and probability of not following social distancing guidelines. This figure plots changes in the predicted probability of non-compliance with 90% confidence intervals based on maximum likelihood estimates. Unadjusted models show the bivariate relationship between the two variables. Adjusted models include the following set of controls: age (with second-order polynomial), gender, education, ideology (with second-order polynomial), inter-personal trust, religion, and self-reported health.

This finding is partly consistent with the pre-registered hypothesis stating that individuals with more extreme political preferences are less likely to comply, likely driven by mechanisms such as lower institutional trust, divided party positioning and associated mass differences in policy preferences. However, our pre-registered hypothesis entails a non-monotonic association between ideology and social distancing (correspondingly, ideology enters the regression in linear and quadratic form) consistent with some prior evidence (Brouard et al. 2020) rather than an asymmetry between those on the left and the right found in our experiment. In the case of the US, our finding is consistent with evidence on polarized elite rhetoric over COVID-19 (Green et al. 2020) and evidence on partisan gaps in social distancing based on mobility patterns from smartphone data, spending behavior, and direct survey question (Allcott et al. 2020; Painter and Qiu 2020). A qualitatively similar pattern exists in New Zealand and Australia, though in Australia it is less pronounced and the confidence intervals of the ideological gap always overlap zero. Notably, the same pattern generally does not show up in the European countries. There, the estimated predictive impact of ideology is approximately null. Following social distancing is not a question of political ideology in the majority of countries under study. In our interpretation, this finding is positive news.

The literature on externalities and social dilemmas (Ostrom 2000) suggests the relevance of interpersonal trust for compliant behavior. An individual’s compliance with non-pharmaceutical interventions contributes to the public good of containing a pandemic. Success depends on a large fraction of people changing their behavior. For an individual, social distancing entails an individual (social) cost but it has positive externalities. However, everybody benefits if the goal is achieved, regardless of whether the person made an effort. In iterated games of social dilemma situations of this type, inter-personal trust is important. If people tend to trust each other, cooperation can be part of an evolutionary stable equilibrium even in the presence of rational egoists. Moreover, compliance with social distancing also has some features of a coordination game, in which people prefer to comply as long as most other people do so as well. The results for Germany are consistent with this trust hypotheses, as people who agree with the statement that “most people can be trusted” are less likely to meet friends and relatives than people who instead say that “You can never be too careful when dealing with other people”. The statistical models suggest that a change from generally not trusting to trusting people is associated with a 17% reduction in the probability of being non-compliant. Again, however, this pattern does not generalize across countries. In most countries there is no statistically significant relationship between inter-personal trust and the following of social distancing rules. In the US the pattern is the opposite (even after adjusting for ideology and other covariates).

Previous work on COVID-19 finds that socio-demographic factors are associated with the (self-reported) willingness to follow social distancing, with gender being one key predictor (Barari et al. 2020; Galasso et al. 2020; Perrotta et al. 2020; Brouard et al. 2020). As shown in the last panel of Figure II, our list experimental estimates are broadly consistent with this. However, they also suggest some qualifications. While women are generally more likely to follow social distancing than men (the estimated unadjusted gender gap is negative for 7 out of 9 countries), this gender difference is not statistically significant in all countries. Moreover, in Germany women were substantively less likely to comply. This result also holds after adjusting for some possible mechanisms, such as labor market participation, ideology, trust, or religion. In Sweden, we find the same pattern but the confidence intervals for the estimates are larger. It is perhaps no coincidence that the two countries with the largest estimated prevalence of not following social distancing (Table I) also exhibit a gender gap with a positive sign.^5^

## Discussion

An important lesson that emerges from our findings is that it may be difficult for policy-makers to learn from other countries’ experiences when crafting policies intended to enhance compliance with public health guidelines. While behavioral social science can draw on a repertoire of experimentally tested ‘nudges’ to enhance compliance (Bavel et al. 2020), our results highlight that the characteristics of individuals less likely to follow health guidelines vary across countries. Thus, behavioral interventions intended to target non-compliers should not be based on the assumption that the characteristics of non-compliers are identical across countries. Rather, the collection of country-specific data would be a better starting point. A somewhat more encouraging aspect of our findings is that while the relevance of political ideology for social distancing is pronounced in some countries—including the US—this is by no means the rule. At least during the lockdown stage of the pandemic, the same was not true in Europe, likely giving policymakers more scope for action in the future.

Our macro-level results illustrate that social distancing appears to be more difficult to maintain when the immediate health impact of the pandemic is comparatively mild and*/*or broader limitations on movement and external sanctions are moderate rather than very strict. While we interpret these results with caution, they underscore the challenge for policymakers trying to open up the economy and society while maintaining behavioral measures that limit the emergence of subsequent outbreaks.

Compared to direct survey questions, the list experimental approach makes our findings less susceptible to measurement errors induced by social desirability bias that have been prominently discussed in the literature. A potential limitation of our experiment is its external validity—understood as the scope of actions captured by the experiment, which focuses on meeting friends or relatives. To study this limitation, we have constructed latent variable models of individuals’ propensity to enact behavioral changes during the pandemic. We analyzed seven direct questions on a range of behaviors, such as more frequent hand washing, or avoiding public places (see OA B). Both a pooled item response theory model and a random coefficients hierarchical factor model, which allows for country-specific response processes, show that a single component explains most of the variation in responses. Reassuringly, there is a strong positive rank correlation between list experimental estimates and country values of the one-dimensional latent factor of behavioral changes (*ρ =* 0.77 and *ρ* = 0.73, respectively, see OA Table B.1). We interpret this as evidence that our evidence from the list experimental taps into a broader underlying behavioral dimension of responses to health guidelines during the pandemic.

## Materials and Methods

### Experimental design

*Implementation:* The list experiment was embedded in a comparative survey conducted via the internet by commercial polling companies in nine countries between April 15-20, 2020.^6^ Data collection was conducted by CSA Research (Australia and the US) and IPSOS (all other countries). OA Table A.1 lists fieldwork periods, sample sizes, and the survey completion rate of participating respondents for each of the nine surveys. Sampling was done as part of existing online panels using quota sampling. The resulting samples were weighted by the survey providers to match Census population margins for gender, age, occupation, region, and degree of urbanization (the latter was not used in New Zealand). All our analyses and descriptive results use probability weights unless otherwise indicated. In total, there are 11,038 respondents. As noted in the pre-analysis plan, the variation in sample sizes across countries reflects resource constraints not related to the list experiment. All participants gave explicit to consent to take part in the survey and answer political questions.

The experiment is conducted by randomly assigning respondent into equal-sized treatment and control groups. Both groups are presented with a list of actions and are asked to report only the sum total of these action performed in the last week. The set of control items includes behavior likely influenced by the pandemic but not violating health guidelines, such as ordering food using online delivery services. The sensitive item presented only in the treatment group states that a respondent met with family or friends who are not part of the same household in the past week. This violates the societal norms in place during the pandemic, and in many countries also violates explicit health advice or orders given by governments. The full wording of the list experiment is given in OA A.2. OA Table A.3 provides an overview of basic individual characteristics for respondents assigned to treatment and control groups.

### Identification

The key assumptions for identification in this experimental design are (i) randomization of treatment (true by design), (ii) no design effects (i.e., responses to control items are not affected by the treatment), (iii) a truthful response to the sensitive item in the treatment condition under the anonymity awarded by the design (respondents are therefore only asked to report sum totals rather than itemized responses). We test possible implication of violating the assumption of no design effects and truthful responses to sensitive item in the treatment group, and generally find no evidence that the design is invalid.

First, a potential problem with the design is that (anticipated) ceiling effects may undermine the anonymity of the response with respect to the sensitive item. A respondent in the treatment group stating that she did all of the listed acts would reveal her norm violation to the researcher and she may thus not respond truthfully. Our set of questions deliberately used innocuous control items that are unlikely to be all answered in the affirmative or all in the negative by most respondents. Data from our experiment show that reported counts (in the control and treatment group) are not concentrated at the ceiling (see OA Table A.2). Furthermore, “self-administration” of the measurement instrument in an online survey context likely reduces non-truthful responses as well (Droitcour et al. 2011: 190).

Second, to make sure to not be associated with the sensitive item, the same individual who reports a non-zero count in the treatment group might want to counterfactually report a zero count in the treatment group. However, inspection of the data from our experiment shows that the share of respondents reporting zero counts is generally not higher under treatment than under control conditions.

Finally, we conducted statistical tests for the assumption of no design effects proposed by Blair and Imai (2012). We generally do not reject the null hypothesis of no design effects (also see OA Table A.2). The UK is an exception if one conducts a test that does not account for multiple-country comparison, but not otherwise. We ensured that all our substantive conclusions are robust to excluding the UK.

### Background

All countries included in the analysis had numerous confirmed COVID-19 cases and all had reported COVID-19 related deaths at the time of the survey around the world (see OA Table A.1). Facing the same pandemic, governments had put in place new health guidelines that emphasized the importance of social distancing to reduce the spread of the virus, alongside other behavioral changes, such as more frequent and thorough hand-washing. Across countries, the general governmental recommendation was not to meet other people and stay home whenever possible. For example, in France all public and private gatherings were banned and in Germany the federal government declared that ‘rule number 1’ was to reduce social contact to a minimum. The US president declared a national emergency on March 13, 2020, and in most US states stay-at-home-orders were in place during the time of the survey (with 94.1% of the population being confined or partially confined according to our data). The exception is Sweden. While Swedish public health authorities also emphasized that everyone has a personal responsibility to prevent transmission and discouraged large events, they did not generally recommend social distancing except for older people.

### Statistical analyses

*Prevalence estimates and macro-level plots:* Under the identifying assumptions listed above, estimates of the prevalence of non-compliance in each country are obtained by simple differences-in-means (using appropriate sampling weights). In the plots of prevalence estimates against macro-level characteristics we add regression lines based on robust regression. In order to capture the uncertainty associated with the estimates of non-compliance, we generate 1,000 replicate data sets via sampling with replacement and estimate the difference in means between treated and control cases in each replicate. We then compute a robust regression using an M-estimator (calculated using iteratively reweighted least squares) with the objective function specified according to Huber (1973: 800) in each data set. Each hairline in Figure I represents one of 1,000 robust regressions; the bold line represents the average regression line.

### Individual-level models

For the individual-level analysis, bivariate and multivariate beta-binomial regression models are used to model the item count in each country as suggested by Imai (2011). Estimates are obtained using maximum likelihood using the Expectation-Maximization algorithm (Blair et al. 2020). The adjusted models reported in Figure II include the following set of individual-level covariates: age (in years) and age squared; an indicator equal to 1 if female, 0 otherwise; an indicator equal to 1 if a respondent has at least a college (BA) degree, 0 otherwise; subjective personal health measured on 5-point scale; the ideological self-placement of the respondent captured using left-right or liberal-conservative 11-point scales (the question reads: “on a scale from 0 to 10, where 0 is left and 10 is right, where would you place yourself politically?”; in the US, the wording is “liberal” and “conservative” rather than “left” and “right”). Ideology is included in linear and quadratic form. We capture interpersonal trust by an indicator variable equal to 1 for respondents who agree with the statement that, generally speaking, “most people can be trusted” (rather than “you can never be too careful when dealing with other people”). Religion is captured by an indictor equal to 1 for respondents who feel close to any religion, 0 otherwise. Missing observations are deleted listwise and the same set of observations are used in the unadjusted and adjusted specification.

## Data Availability

Survey data and code files will be deposited on Harvard Dataverse upon publication.

## Online Appendix to

### A. Experiment details

#### A.1. Survey fieldwork

The surveys were in the field between April 15 and April 20 2020 carried out by IPSOS in Austria, France, Germany, Italy, New Zealand, Sweden, and the United Kingdom. In Australia and the United States data collection was conducted by CSA Research. Table A.1 lists fieldwork periods, sample sizes, and the survey completion rate of participating respondents in 9 surveys. The last two columns show the macro variables employed in our plot, the median number of deaths ascribed to Covid-19 in the week prior to the survey as share of the total population, and an index of the stringency of the overall governmental response from the Oxford Covid-19 Government Response Tracker.

Sampling was done as part of ongoing online panels using quota sampling. Dropout rates are relatively low. In most countries more than 90% of respondents completed the survey after agreeing to participate. In Australia and the United States, the completion rate is closer to 70%. The resulting samples were weighted by the survey providers to match Census population margins for gender, age, occupation, region, and degree of urbanization (the latter was not used in New Zealand). We excluded 32 cases (0.266%) with excessively large weights (*>* 5).^1^ All our analyses and descriptive results use probability weights unless otherwise indicated.

#### A.2. List experiment question wording

The list experiment is introduced by an identical statement for treated and control cases. Respondents are prompted with “How many of these things have you done last week? You do not need to tell me which ones you have done, just how many.”^2^ Participants were randomly assigned to two groups; 50% of participants received list A, 50% received list B.

- LIST A [Control group]
  1. I went to the doctor or to the hospital.
  2. I used public transportation to get to work.
  3. I exercised outdoors.
  4. I ordered food using an online delivery service.
- LIST B [treatment group]

**Table A.1.** Survey details.

|  | Fieldwork | Sample size | Resp. rate<sup>a</sup> | Completion rate<sup>a</sup> | Deaths<sup>b</sup> | Gov. response index<sup>c</sup> |
| --- | --- | --- | --- | --- | --- | --- |
| Australia | 04/15 - 04/19 | 1 007 | 0.10 | 0.76 | 3 | 73.2 |
| Austria | 04/15 - 04/18 | 1 000 | 0.33 | 0.95 | 49 | 81.5 |
| France | 04/15 - 04/16 | 2 020 | 0.47 | 0.96 | 256 | 90.7 |
| Germany | 04/16 - 04/18 | 2 000 | 0.31 | 0.93 | 54 | 73.2 |
| Italy | 04/15 - 04/17 | 997 | 0.37 | 0.94 | 367 | 93.5 |
| New Zealand | 04/15 - 04/18 | 998 | 0.38 | 0.94 | 2 | 96.3 |
| Sweden | 04/16 - 04/18 | 1 009 | 0.33 | 0.95 | 118 | 38.0 |
| United Kingdom | 04/15 - 04/17 | 1 000 | 0.35 | 0.94 | 224 | 75.9 |
| United States | 04/15 - 04/20 | 2 007 | 0.12 | 0.68 | 114 | 74.5 |

<sup>a</sup> Response rate $S/I$ , completion rate $C/(S-Q)$ ; $I$ is the number of individuals invited, $S$ the number of started surveys, $Q$ number of surveys removed due to quota being fulfilled, $C$ number of completed surveys.
<sup>b</sup> Median number of deaths per million inhabitants in week prior to survey. Source: COVID-19 Data Repository, Center for Systems Science and Engineering, JHU (Dong et al. 2020).
<sup>c</sup> Government response stringency index. Source: Oxford Covid-19 Government Response Tracker (Hale et al. 2020).

1. I went to the doctor or to the hospital.
2. I used public transportation to get to work.
3. I exercised outdoors.
4. I met with two or more friends or relatives who do not live with me.
5. I ordered food using an online delivery service.

#### A.3. Exploring experimental design assumptions

The first two columns of Table A.2 shows average item counts in the control group (as well as the coefficient of variation) by country. They indicate that ceiling effects are not a likely concern. In all countries the control group mean item count is below 1.5 with a coefficient of variation around one. However, observing responses close to zero raises the potential issue that a large fraction of respondents choose the rational strategy of replying with ‘0’ simply to ensure that there is no chance that they can be associated with a social norm violation. Column *Y*_0_ and *Y*_1_ of Table A.2 reports the fraction of respondents reporting having committed none of the acts in the list presented to them for the control and treatment group, respectively. If many respondents indeed follow a rational ‘0’ strategy, we would expect to find that the fraction of ‘0’ responses to be considerably higher in the treated group (who do see the norm violation item) than in to the control group. But, while we do find a seizable share of ‘0’ respondents in the control group, the corresponding share in the treatment group is generally the same or lower. These results suggest that those exposed to the norm violation treatment are not more likely to shift to a strategy of ‘0’ responses. The exception to this pattern is the United Kingdom, where we find that the fraction of ‘0’ responses among the treated is 6 percentage points higher than among the control group.

**Table A.2.** **Size of control and treatment group, item counts in control group (means and coefficients of variation), proportion of zeros in control and treatment group, and test for design effect**.

| Country | $N_0$ | $N_1$ | $Y_0$ avg. | $Y_0$ CV | Prop. zeros | | Design effect | |
| --- | --- | --- | --- | --- | --- | --- | --- | --- |
| | | | | | $Y_0$ | $Y_1$ | $p$ | $p_{BH}$ |
| Australia | 513 | 494 | 1.395 | 0.823 | 0.23 | 0.22 | 1.000 | 1.000 |
| Austria | 498 | 498 | 1.061 | 1.014 | 0.37 | 0.27 | 0.915 | 1.000 |
| France | 1014 | 1006 | 0.696 | 1.187 | 0.48 | 0.45 | 0.984 | 1.000 |
| Germany | 1000 | 1000 | 1.081 | 1.009 | 0.36 | 0.12 | 0.098 | 0.443 |
| Italy | 498 | 499 | 0.753 | 1.318 | 0.53 | 0.54 | 0.616 | 1.000 |
| New Zealand | 500 | 498 | 1.131 | 0.681 | 0.16 | 0.15 | 1.000 | 1.000 |
| Sweden | 502 | 507 | 0.912 | 1.148 | 0.42 | 0.29 | 1.000 | 1.000 |
| United Kingdom | 500 | 500 | 1.244 | 0.757 | 0.18 | 0.24 | 0.021 | 0.192 |
| United States | 956 | 1013 | 1.121 | 1.001 | 0.36 | 0.32 | 1.000 | 1.000 |

*Note:* Means and proportions weighted by sample-inclusion probability. Last two columns show null-hypothesis tests of the no design effect assumption proposed by Blair and Imai (2012: sec. 3.1). $p_{BH}$ denotes $p$ -values additionally adjusted for false-discovery rates of multiple-country comparisons using the Benjamini-Hochberg procedure (with $\alpha = 0.05$ ).

Blair and Imai (2012) provide a more sophisticated test of possible design effects in list experiments. A design effect occurs when responses to the control items change due to the presence of the norm violating item. This might be due to respondents evaluating items relative to each other, emotional responses induced by the presence of a sensitive item, or the rational ‘0’ strategy discussed above. The final two columns of Table A.2 shows *p* values for tests of the null hypothesis of no design effect. The column labelled *p*_*BH*_ additionally adjusts *p* values for multiple country tests using the false-discovery rate controlling procedure of BH.^3^ The results clearly do not indicate the presence of design effects in 8 out of 9 experiments: we cannot reject the null hypothesis of no design effect in all countries except the United Kingdom. In the United Kingdom the statistical detection of design effects depends on the decision to adjust for multiple comparisons. Thus, results for the UK should at least be treated with caution. We therefore ensured that excluding the United Kingdom does not affect our substantive conclusions (note that only our macro plot in Figure I pools information from different countries).

#### A.4. Sample characteristics at baseline for treatment and control units

Table A.3 provides an overview of basic individual characteristics for respondents assigned to treatment and control groups. For each, the first column displays means followed by the standard error of the mean. The third column indicates the sample standard deviation. The final column lists the difference in means between treated and control groups. While randomization guarantees balance on covariates in expectation, we also find that observable characteristics in our sample are fairly balanced between treatment and control groups. Slightly more noticeable differences emerge for average ages in France, New Zealand, and Sweden, where members of the treatment group are about one to two years older. We do provide specifications that account for age differences in our estimates of individual-level determinants of non-compliance.

**Table A.3.** Covariates at baseline.

|  | Control |  |  | Treated |  |  | Diff<br>(4-1) |
| --- | --- | --- | --- | --- | --- | --- | --- |
|  | Mean<br>(1) | s.e.<br>(2) | s.d.<br>(3) | Mean<br>(4) | s.e.<br>(5) | s.d.<br>(6) |  |
| *Australia* |  |  |  |  |  |  |  |
| Age | 45.33 | 0.63 | 14.36 | 45.29 | 0.65 | 14.47 | −0.040 |
| Female | 0.49 | 0.02 | 0.50 | 0.49 | 0.02 | 0.50 | 0.001 |
| Ideology | 5.39 | 0.10 | 2.13 | 5.73 | 0.11 | 2.16 | 0.341 |
| Trust | 0.53 | 0.02 | 0.50 | 0.50 | 0.02 | 0.50 | −0.028 |
| *Austria* |  |  |  |  |  |  |  |
| Age | 47.45 | 0.77 | 17.29 | 46.63 | 0.72 | 16.03 | −0.822 |
| Female | 0.50 | 0.02 | 0.50 | 0.53 | 0.02 | 0.50 | 0.036 |
| Ideology | 5.03 | 0.09 | 2.04 | 4.93 | 0.09 | 2.05 | −0.098 |
| Trust | 0.47 | 0.02 | 0.50 | 0.47 | 0.02 | 0.50 | −0.005 |
| *Germany* |  |  |  |  |  |  |  |
| Age | 48.49 | 0.52 | 16.57 | 49.30 | 0.52 | 16.56 | 0.803 |
| Female | 0.53 | 0.02 | 0.50 | 0.49 | 0.02 | 0.50 | −0.038 |
| Ideology | 4.73 | 0.06 | 1.90 | 4.71 | 0.06 | 1.99 | −0.022 |
| Trust | 0.48 | 0.02 | 0.50 | 0.50 | 0.02 | 0.50 | 0.024 |
| *France* |  |  |  |  |  |  |  |
| Age | 49.23 | 0.51 | 16.27 | 50.84 | 0.50 | 15.74 | 1.610 |
| Female | 0.54 | 0.02 | 0.50 | 0.51 | 0.02 | 0.50 | −0.025 |
| Ideology | 5.42 | 0.08 | 2.34 | 5.20 | 0.08 | 2.35 | −0.218 |
| Trust | 0.36 | 0.02 | 0.48 | 0.39 | 0.02 | 0.49 | 0.027 |
| *United Kingdom* |  |  |  |  |  |  |  |
| Age | 46.54 | 0.72 | 16.09 | 46.56 | 0.73 | 16.37 | 0.019 |
| Female | 0.50 | 0.02 | 0.50 | 0.53 | 0.02 | 0.50 | 0.033 |
| Ideology | 4.91 | 0.11 | 2.30 | 4.86 | 0.10 | 2.14 | −0.048 |
| Trust | 0.50 | 0.02 | 0.50 | 0.51 | 0.02 | 0.50 | 0.012 |
| *Italy* |  |  |  |  |  |  |  |
| Age | 48.84 | 0.76 | 17.05 | 49.47 | 0.75 | 16.65 | 0.637 |
| Female | 0.52 | 0.02 | 0.50 | 0.52 | 0.02 | 0.50 | 0.004 |
| Ideology | 4.85 | 0.11 | 2.37 | 5.20 | 0.12 | 2.60 | 0.354 |
| Trust | 0.34 | 0.02 | 0.47 | 0.36 | 0.02 | 0.48 | 0.022 |
| *New Zealand* |  |  |  |  |  |  |  |
| Age | 45.34 | 0.72 | 16.14 | 47.72 | 0.78 | 17.42 | 2.376 |
| Female | 0.52 | 0.02 | 0.50 | 0.52 | 0.02 | 0.50 | −0.005 |
| Ideology | 5.06 | 0.11 | 2.26 | 5.09 | 0.11 | 2.21 | 0.034 |
| Trust | 0.58 | 0.02 | 0.49 | 0.59 | 0.02 | 0.49 | 0.012 |
| *Sweden* |  |  |  |  |  |  |  |
| Age | 48.06 | 0.72 | 16.18 | 49.15 | 0.73 | 16.43 | 1.097 |
| Female | 0.53 | 0.02 | 0.50 | 0.48 | 0.02 | 0.50 | −0.044 |
| Ideology | 5.13 | 0.12 | 2.51 | 5.13 | 0.12 | 2.64 | 0.002 |
| Trust | 0.58 | 0.02 | 0.49 | 0.54 | 0.02 | 0.50 | −0.033 |
| *United States* |  |  |  |  |  |  |  |
| Age | 46.11 | 0.55 | 17.28 | 45.87 | 0.54 | 17.21 | −0.233 |
| Female | 0.51 | 0.02 | 0.50 | 0.50 | 0.02 | 0.50 | −0.007 |
| Ideology | 5.70 | 0.09 | 2.75 | 5.38 | 0.09 | 2.93 | −0.329 |
| Trust | 0.45 | 0.02 | 0.50 | 0.43 | 0.02 | 0.50 | −0.021 |

#### A.5. Existing social ties

As noted in the main text, Figure A.1 plots the share of the population in each country not following social distancing, estimated from the list experiment, against pre-COVID-19 social connections measured by the average time spent socializing with friends and family. Specifically, we use data from the OECD (2020: Figure 11.3) on time (in hours) spent per week interacting with family and friends as a primary activity calculated from Eurostat’s Harmonised European Time Use Surveys (from 2018 or previous years). Figure A.1 illustrates that pre-pandemic patterns of socializing are not strongly related to the share of individuals not following health guidelines during the pandemic. The Spearman rank correlation of socializing and non-compliance with social distancing is 0.02 with a *p*-value of 0.98.

**Figure A.1.**
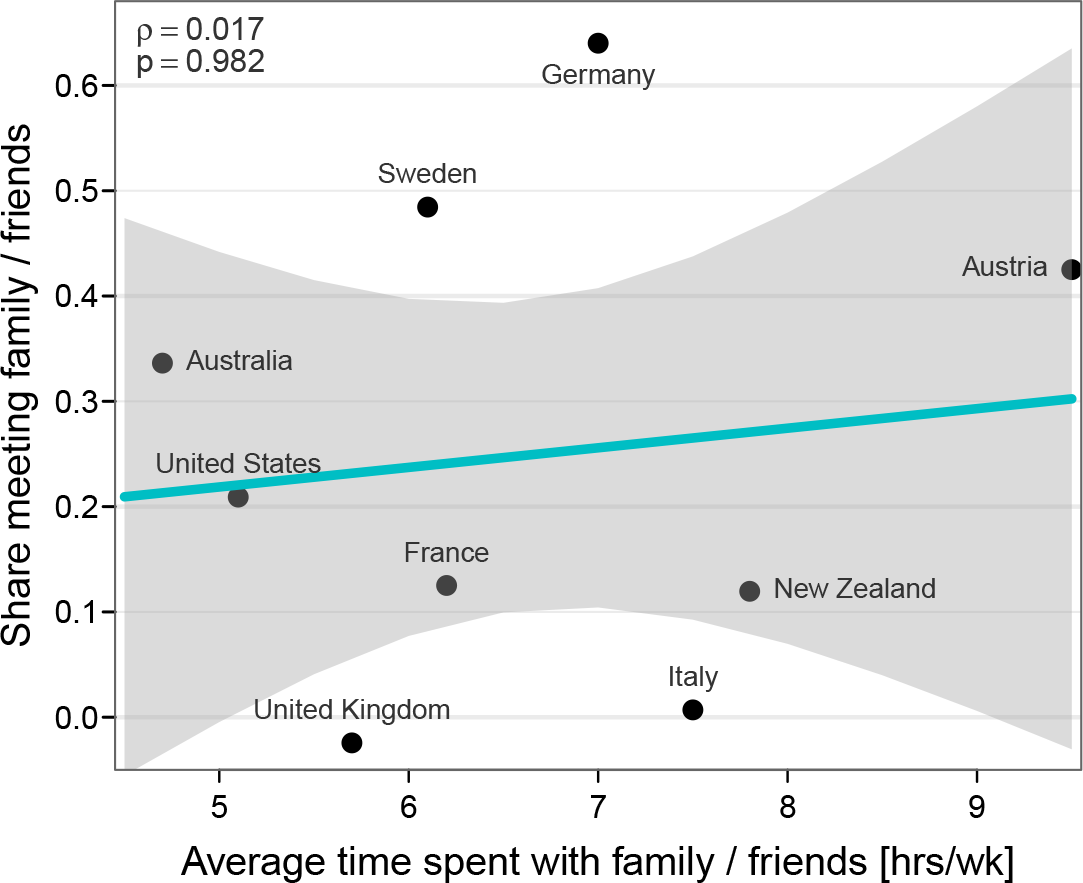
Social ties and prevalence of noncompliance. This figure shows that existing patterns of social interactions are not strongly related to the share of individuals not following health guidelines during the pandemic. It plots the average time spent socializing with family or friends [as primary activity, in hours/week] around 2018 (data from OECD 2020) against experimental estimates of the share of individuals meetings family or friends despite health guidelines in 2020. Robust regression line with confidence bands superimposed. The Spearman rank correlation between both measures is 0.017 with a *p*-value of 0.982.

### B. Latent variable model of items measuring behavioral changes

In this section we describe the construction of a one-dimensional latent factor capturing behavioral changes following the pandemic more broadly than our list experiment.

Our survey contains a battery of items asking respondents if they have changed their behavior since the beginning of the pandemic. These were placed distantly after the survey experiment. They are presented with a list of items:

- washing your hands more often and/or for a longer amount
- coughing or sneezing into your elbow or a tissue
- stopped greeting others by shaking hands, hugging or kissing
- keep a distance of [six feet] between yourself and other people^4^
- reduced your trips outside home
- avoid busy places (public transportation, restaurants, sport)
- stopped seeing friends

Responses for each item were originally recorded on an 11-point response scale. This question format produces extreme skewness of responses: for many items more than 50% of respondents chose the highest two out of 11 categories. We dichotomized all items such that responses other than ‘9’ and ‘10’ (the highest two categories) indicate that respondents likely did not adjust their behavior (or only did so in a selective manner).

We first study if the configuration of these items follows similar patterns in each country and if they can be summarized by a low-dimensional vector of latent variables. Figure B.1 summarized results from a nonlinear principal components analysis (Gifi 1990) of our dichotomized item battery estimated separately in each country. Panel **A** shows the eigenvalues for seven principles components in each country. It suggest that one component captures a dominant share of variation in each country. All eigenvalues for components other then the first are less then 1 (save for Sweden, which is barely above 1 for component 2). Similarly, panel **B**, which plots component loadings for each item on the first two principal components for each country, suggests that the predominant variation takes place on the first component. Based on this initial exploratory analysis, we specify one-dimensional latent factor/IRT models described next.

#### B.1. Pooled IRT model and issues of measurement equivalence

A simple first latent variable model for these items is a standard two-parameter IRT model estimated on the pooled sample. The parameters of this model are item intercepts, *τ*, (referred to as “difficulties” in the IRT literature) and coefficients, *λ*, capturing how a unit increase in the latent variable shifts the propensity of observing each item (“discrimination parameters”). Expressed briefly, and using factor-analytic notation (Takane and de Leeuw 1987), for any given item *y* the the model takes the form *y*_*i*_ = *τ* + *λf*_*i*_ + *εi*, where the distribution of residuals *ε* is normal with variance fixed to 1 and the latent variable *f* is distributed normally with mean 0 and unit standard deviation (for a detailed introduction to IRT models see, e.g., van der Linden 2016; Hambleton et al. 1991). Estimating this model in a Bayesian framework using the Gibbs sampler, draws from the posterior distribution of *f* can be obtained straightforwardly and aggregated to country-specific averages.

**Figure B.1.**
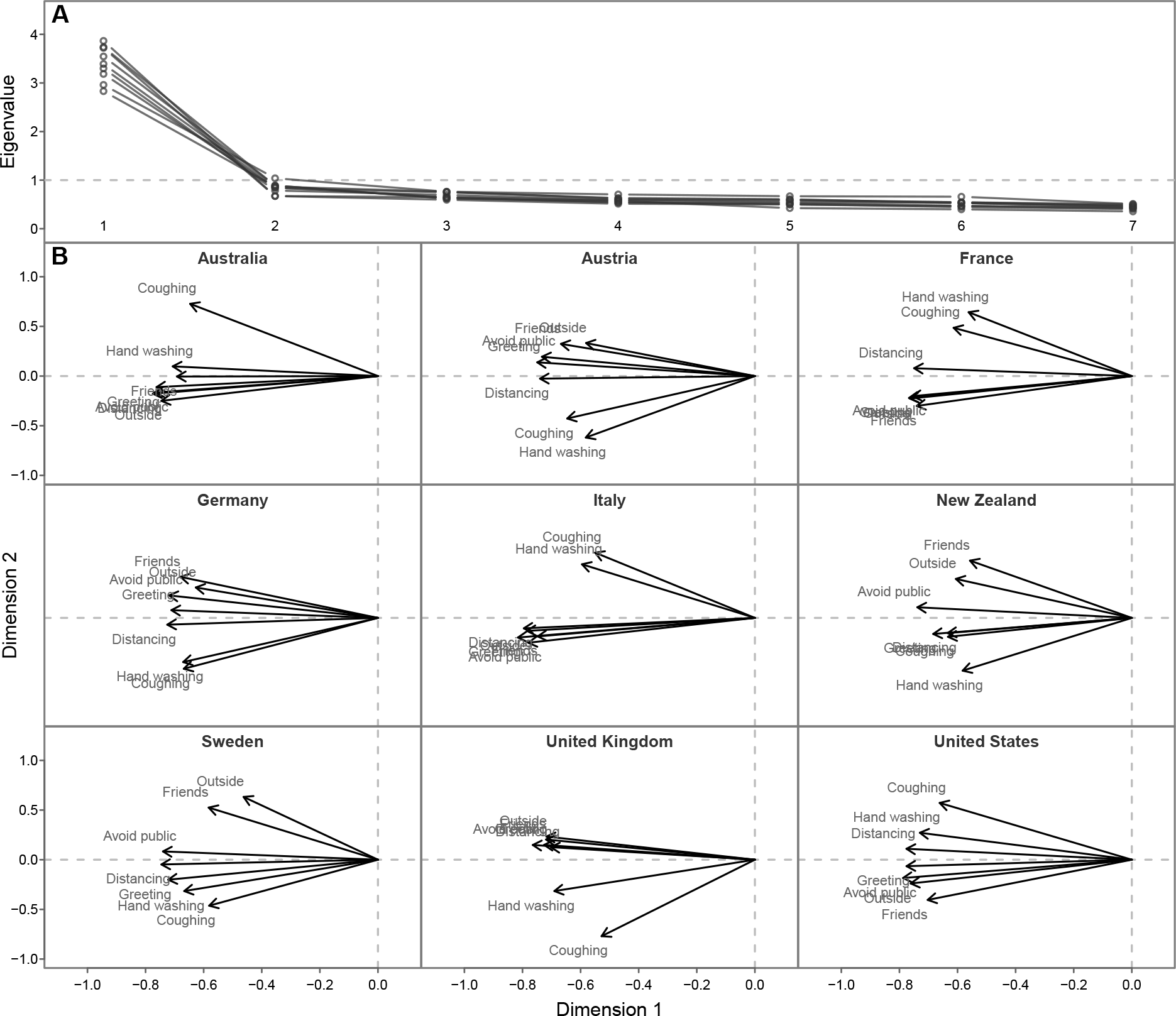
Nonlinear principal components analysis of behavioral adjustment battery. Panel **A** shows the eigenvalues of seven principal components for each of 9 countries. It indicates that extracting one component captures a large proportion of total variation. Panel **B** shows component loadings plot for the first two largest components in each country. The configuration of the loadings in each country also suggests that a one-dimensional factor captures the most important differences between respondents.

The pooled model ignores the problem of measurement equivalence. Pooling information from different countries with potentially heterogenous response processes might make it invalid to compare means of the latent factor (see, e.g., Davidov et al. 2014; Stegmueller 2011). The factor analytic literature usually distinguishes between different degrees of measurement invariance (e.g. Millsap 2011): *configural invariance* assumes a similar fundamental factor structure in each country (as emerged in our PRINCALS analysis above), but puts no equality restrictions on any model parameters in different countries. *Metric invariance* adds equality constraints for loadings, while *scalar invariance* adds equality constrains for both loadings and intercepts. Under essential country heterogeneity in response processes, factor means and variances are only identified under the scalar invariance restriction. Thus, imposing equality in loadings and intercepts in the pooled model where it does not exist leads to distorted estimates of the latent factor and the resulting country means are not quantitatively comparable.

#### B.2. Random coefficient hierarchical factor model

We estimate an alternative latent variable model that explicitly allows for country-differences in differential item functioning following the proposals by de Jong et al. (2007) and Fox and Verhagen (2010). They key idea is to specify a hierarchical factor model with random coefficients (Ansari et al. 2000, 2002) allowing for heterogeneity in item parameters while being anchored to a common mean.

Denote by *y*_*i jk*_ the response of person *i* (*i* = 1, … *N*_*j*_) in country *j* (*j* = 1, … *J* = 9) to survey item *k* (*k* = 1, …, *K* = 7) probing if he or she changed health-relevant behaviors. Each item is specified as a probit equation and we work with the underlying latent variables 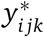, which are available via data augmentation during the Gibbs sampler (Albert and Chib 1993). We specify each 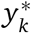 as being driven by an underlying latent factor *f*_*W*_. We estimate the following measurement system

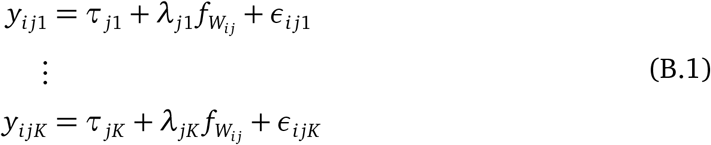

where *τ* are item intercepts, *λ* are factor loadings and 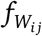 is a latent factor representing individual propensity to change behavior. For identification, 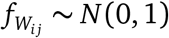 as in standard IRT models.^5^ Residuals *εi jk* are also called uniqenesses in the factor analysis literature and are assumed independent after conditioning on the latent trait and distributed mean zero with unit variance (in order to fix the underlying variance of the probit model). Both item intercepts and loadings are free to vary over countries and are anchored by the following hierarchical factor structure:

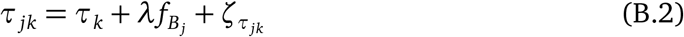

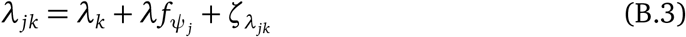

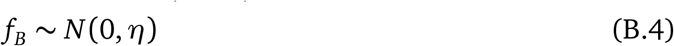

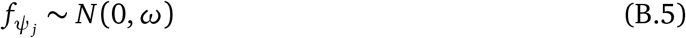

where random item effects are distributed 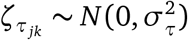 and 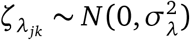.

Fox (2010) discusses the identification constraints needed to separately identify varying factor means and variances with both intercepts and leadings hierarchically modeled. We follow the strategy outlined in Asparouhov and Muthén (2015). Note that the loadings *λ* are equal for 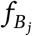 and 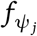.

The variation in item intercepts over countries is captured by 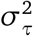 while the variation in loadings is captured by 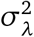. The systematic country-variation of individual factor means is captured by *η*; the variation in the factor variance is captured by *ω*. Using posterior draws from 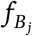 we can straightforwardly obtain our country-level estimate of health guideline related behavior.^6^

#### B.3. Resulting estimates and comparison to list experiment

Table B.1 shows latent factor estimates for each country obtained using both modeling approaches compared to the estimates from our list experiment. The scaling of both quantities makes numerical comparisons difficult: the latent variable is normalized to have mean 0 (with a fixed standard deviation of 1) while estimates from the list experiment lie in the unit interval. However, comparing the rank order of estimates reveals that estimates from the list experiment follow a pattern comparable to estimates from the pooled latent variable model, which captures a much broader range of pandemic-related behavioral changes. Sweden, Australia, and Germany show the largest factor estimates and are also among the four largest experimental estimates (with the exception of Austria). The United Kingdom and Italy, both with list experimental estimates of essentially zero also emerge among the bottom three countries ranked via the latent factor estimates. The rank correlation between both sets of estimates is 0.77 with an exact *p*-value of 0.021. This pattern is replicated using the more flexible random coefficient factor model (with 31 estimated parameters) shown in the final column of Table B.1. The rank correlation between experimental estimates and the latent country-level factor *f*_*B*_ is 0.73 with a *p*-value of 0.031.

**Table B.1.**
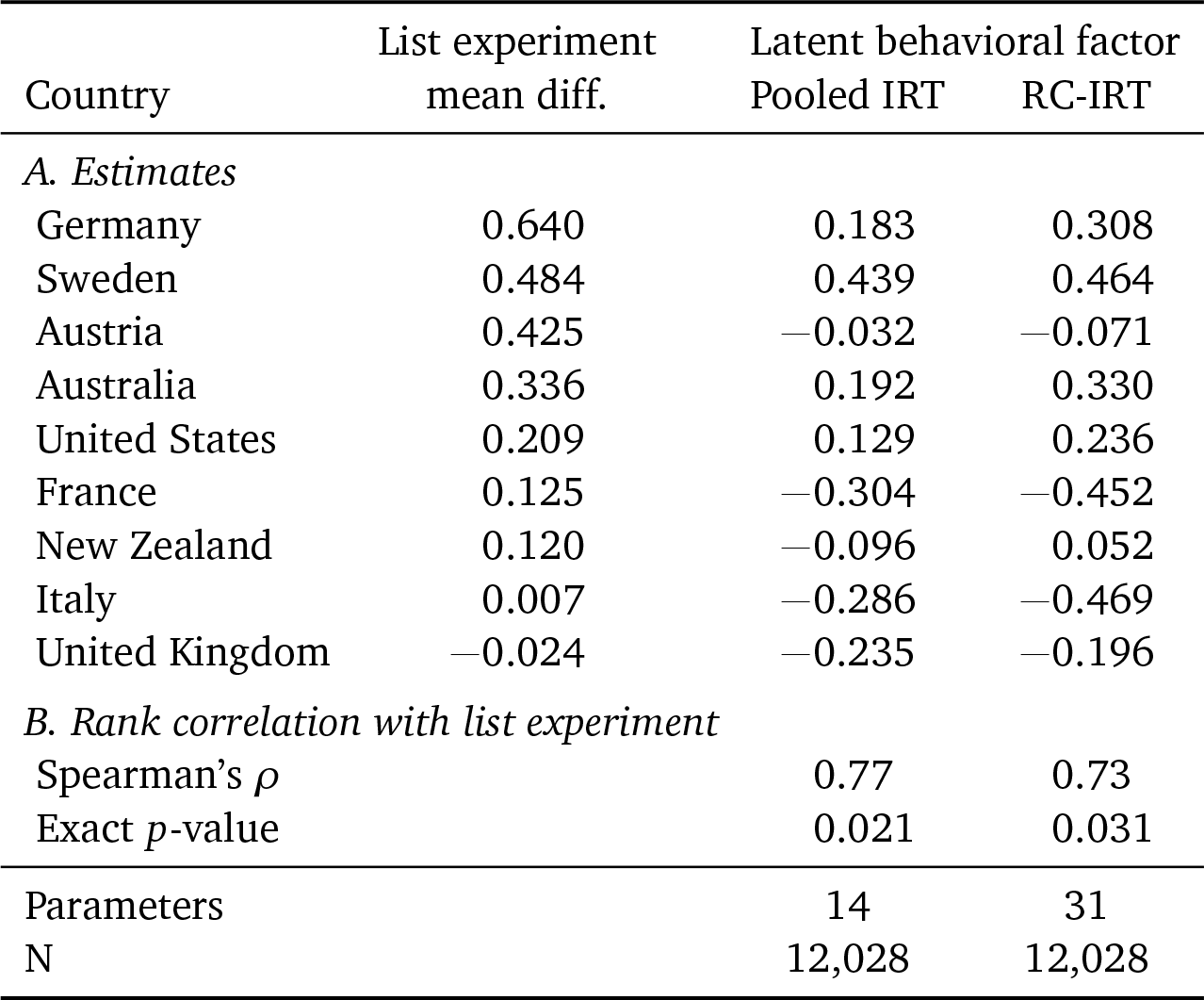
**Relationship between experimental estimates and country values of one-dimensional latent factor of health guidance behavioral adjustments**.

## Footnotes

1 The list experimental approach has become very popular recently, and has been employed across disciplines, from studying substance abuse, HIV risk behavior, employee theft, to brand preferences (see Blair and Imai 2012 for a list of applications). In political science it has been used, among others, to study turnout, racial prejudice, or support for abortion (Kuklinski et al. 1997; Holbrook and Krosnick 2010; Rosenfeld et al. 2016). With respect to compliance with social distancing measures during the COVID-19 pandemic, country studies have tackled the problem of survey response bias using either “face-saving” strategies that allow respondents to rationalize non-compliant behavior (Daoust et al. 2020) or single-country list experiments (Larsen et al. 2020; Munzert and Selb 2020). Results of existing studies are incongruent (finding varying degrees of non-compliance using different measurement tools) and call for a comparative study using a unified design.

2 While reporting standards vary across countries, these data have been widely reported in the media and thus shaped the public salience of the pandemic and its associated risks.

3 OA Table A.1 shows country values of both the macro variables.

4 The anonymous pre-analysis plan is available at http://aspredicted.org/blind.php?x=hv7yv2

5 While we cannot address this possibility with our data, one hypothesis for future research is once the social equilibrium is to take social distancing less seriously, gender roles (e.g., taking care of dependents) may entail less social distancing for women than men.

6 The data were collected for the collaborative project “Citizens’ Attitudes Under COVID-19 Pandemic” by the following research team: Sylvain Brouard (Sciences Po, CEVIPOF & LIEPP), Michael Becher (IAST-Université Toulouse Capitole 1), Martial Foucault (Sciences Po-CEVIPOF), Pavlos Vasilopoulos (University of York), Vincenzo Galasso (Bocconi University), Christoph Hönnige (University of Hanover), Eric Kerrouche (Sciences Po-CEVIPOF), Vincent Pons (Harvard Business School), Hanspeter Kriesi (EUI), Richard Nadeau (University of Montreal), Dominique Reynié (Sciences Po-CEVIPOF), Daniel Stegmueller (Duke University).

1 Results have been replicated with these cases included as well.

2 French, German, Italian and Swedish language versions of these item lists are available upon request.

3 Note that the Blair Imai test already Bonferroni-adjusts *p* values for multiple testing *within* countries (Blair and Imai 2012: 64).

4 The distance used in this item corresponds to the health guidelines of each country at the time: 6 feet in New Zealand, UK, US; 3 feet in Australia, 1m in Austria, France, Italy; 2m in Germany; 1.5m in Sweden.

5 The sign of the latent variable is not identified (Anderson and Rubin 1956). In our application this is of no concern since its orientation (“less” inclined to follow health guidance) is easily established from the pattern of loadings.

6 The model is estimated using Gibbs sampling using latent data augmentation for the dichotomous variables. We specify normal priors for all *λ* and *τ* with mean 0 and prior variance 10. Random effect variance terms are given inverse Gamma priors with shape and scale set to 0.001. The prior for covariance matrix of the two factor variances *η* and *ω* is inverse Wishart with *V* = *diag*(1) and degrees of freedom set to *v* = *dim*(*V*) + 1 = 3.

## Notes

### Competing Interest Statement

The authors have declared no competing interest.

### Funding Statement

Becher acknowledges IAST funding from the French National Research Agency (ANR) under the Investments for the Future (Investissements d'Avenir) program, grant ANR-17-EURE-0010. Brouard acknowledges the financial support from ANR - REPEAT grant (Special COVID-19), CNRS, Fondation de l'innovation politique, regions Nouvelle-Aquitaine and Occitanie. Stegmueller's research was supported by the National Research Foundation of Korea (NRF-2017S1A3A2066657).

### Author Declarations

The Review Board for Ethical Standards in Research at the Toulouse School of Economics and the Institute for Advanced Study approved the study (ref.code 2020-04-001).

